# Design of effective outpatient sentinel surveillance for COVID-19 decision-making: a modeling study

**DOI:** 10.1101/2022.10.21.22281330

**Authors:** Kok Ben Toh, Manuela Runge, Reese AK Richardson, Thomas J Hladish, Jaline Gerardin

## Abstract

**Background:** Decision-makers impose COVID-19 mitigations based on public health indicators such as reported cases, which are sensitive to fluctuations in supply and demand for diagnostic testing, and hospital admissions, which lag infections by up to two weeks. Imposing mitigations too early has unnecessary economic costs, while imposing too late leads to uncontrolled epidemics with unnecessary cases and deaths. Sentinel surveillance of recently-symptomatic individuals in outpatient testing sites may overcome biases and lags in conventional indicators, but the minimal outpatient sentinel surveillance system needed for reliable trend estimation remains unknown.

**Methods:** We used a stochastic, compartmental transmission model to evaluate the performance of various surveillance indicators at reliably triggering an alarm in response to, but not before, a step increase in transmission of SARS-CoV-2. The surveillance indicators included hospital admissions, hospital occupancy, and sentinel cases with varying levels of sampling effort capturing 5, 10, 20, 50 or 100% of incident mild cases. We tested 3 levels of transmission increase, 3 population sizes, and condition of either simultaneous transmission increase, or lagged increase in older population. We compared the indicators’ performance at triggering alarm soon after, but not prior, to the transmission increase.

**Results:** Compared to surveillance based on hospital admissions, outpatient sentinel surveillance that captured at least 20% of incident mild cases could trigger alarm 2 to 5 days earlier for a mild increase in transmission and 6 days earlier for moderate or strong increase. Sentinel surveillance triggered fewer false alarms and averted more deaths per day spent in mitigation. When transmission increase in older populations lagged increase in younger populations by 14 days, sentinel surveillance extended its lead time over hospital admissions by an additional 2 days.

**Conclusions:** Sentinel surveillance of mild symptomatic cases can provide more timely and reliable information on changes in transmission to inform decision-makers in an epidemic like COVID-19.

## Background

COVID-19 is a contagious disease with more than 6 million deaths reported within two years of the global pandemic starting in early 2020 (1,2). Prior to widespread availability of vaccines, policy makers relied on non-pharmaceutical interventions or mitigative actions, such as lockdown and business restrictions, to curb the spread of SARS-CoV-2 (3–7). These actions “flatten the curve”, allowing governments to reduce the COVID-19 burden to the healthcare system, to maintain the quality of care, and to save lives. Timely introduction of interventions is important because imposing mitigations too early has unnecessary economic costs, while imposing too late often fails to control the epidemics and leads to many preventable cases and deaths. The decision criteria for imposing mitigations are often based on public health indicators such as reported cases and hospital admissions (8).

These commonly-used indicators have limitations when used to estimate transmission trends. Cases can be biased because they are sensitive to fluctuations in supply and demand for diagnostic testing (9–11). Because the data needed to adjust for this bias is unavailable, cases often provide unreliable approximation of the transmission trend. Although hospital admission data are less sensitive to fluctuations in testing demands, hospital admissions lag infection by up to two weeks (12–16). Decisions made based on trends in hospital admissions thus may result in delayed action to mitigate an incoming epidemic wave. Furthermore, most COVID-19 patients do not require hospitalization, making admissions data prone to high variability due to small numbers and reducing its suitability for decision-making.

Sentinel surveillance of recently-symptomatic people in outpatient testing sites could overcome these biases and lags (17). Under outpatient sentinel surveillance, symptom status, symptom onset, and date of testing site visit are recorded for each case. Sentinel cases here are defined as recently symptomatic people with symptom onset within 4 days of testing. By focusing on outpatient symptomatic cases, the influence of asymptomatic testing is removed, and the impact of fluctuations in test availability reduced, each of which can introduce selection bias. With known symptom onset dates, we can remove care-seeking and reporting delays, and more accurately infer the infection time series and trends in transmission.

Compared to hospital admissions, sentinel cases have a shorter delay between the point of infection and detection by the surveillance system. In the event of an increase in transmission, sentinel cases should thus provide a more timely alarm and enable earlier action against a new epidemic wave. The city of Chicago, USA, evaluated this surveillance scheme during the COVID-19 pandemic. Even with few participating testing sites, the Chicago Department of Public Health found that sentinel cases were indeed more timely than hospital admissions in estimating transmission trends (17). However, the estimated trends were less certain due to low sampling effort, defined as the proportion of incident symptomatic infections captured in the surveillance system.

Understanding the effects of sampling effort is critical in evaluating the potential feasibility and benefits of using sentinel surveillance to guide decision-making. Moreover, because hospitalization rates are lower in younger populations, sentinel surveillance may be especially useful when transmission surges in younger populations first, before it surges in older populations, as it had been observed in several epidemic waves in the USA and UK (18,19).

Mathematical transmission models can provide insight into the potential benefits of an outpatient sentinel surveillance system by assessing the effects of sampling effort. Mechanistic models have been used to understand disease dynamics, forecast hospital needs, and evaluate intervention scenarios throughout the COVID-19 pandemic (20–25). Modeling has been used to inform decision-makers about optimal lockdown, reopening and mitigation strategies (26–29). While mathematical models have been used to compare testing strategies (26,30–32), few modeling studies have been conducted to compare surveillance designs for timely and appropriate imposition of COVID-19 mitigations (33).

To characterize the minimal sampling effort for which an outpatient sentinel surveillance system needed for reliable estimation of trends in transmission, we use a stochastic compartmental model of SARS-CoV-2 transmission to evaluate the performance of various surveillance indicators and sampling efforts at reliably triggering an alarm in response to, but not before, a step increase in transmission rate.

## Methods

### SEIR model and simulation framework overview

We used a published stochastic SEIR compartmental model (33) to simulate SARS-CoV-2 transmission and COVID-19 disease states (Figure 1A). The model included multiple symptom statuses (asymptomatic, presymptomatic, mild, and severe), and multiple severe disease outcomes (requiring hospitalization, critical illness requiring intensive care unit (ICU) admission, and deaths). We simplified the model by assuming that only severely ill individuals would isolate, and that isolation led to lowered infectiousness.

**Figure 1.**
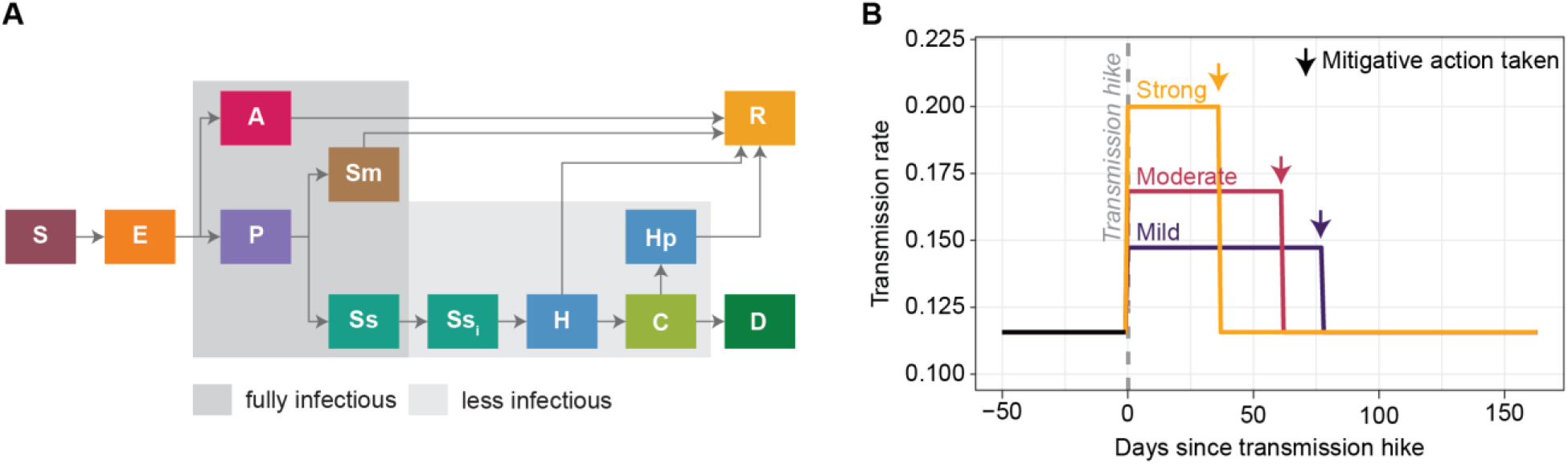
Model framework used in this study. (A) SEIR model structure to simulate SARS-CoV-2 transmission. S=Susceptible, E=Exposed, A=Asymptomatic, P=Presymptomatic, Sm=Mild symptomatic, Ss=Severe symptomatic, H=Requires hospitalization, C=Critically ill (ICU), Hp=Hospitalization post-ICU, D=Death, R=Recovered. All severely symptomatic persons were isolated (SS_i_) few days after entering the state and become less infectious. (B) Simulated transmission rate begins low and steady, then experiences a step increase (transmission hike) at 3 possible magnitudes to create different strengths of epidemic waves. When the surveillance indicator reaches a predetermined threshold, mitigative actions are triggered in the simulation and the transmission rate is decreased to its baseline rate.

We use the model parameters from the previous study (33) which set the hospital and ICU lengths-of-stay based on data from Chicago and from literature during the first COVID-19 wave beginning March 2020 (34,35). This previous study also fitted a time-varying transmission rate to daily ICU census, hospital census, and reported deaths in Chicago. See SI for details of parameterization.

The original model was simulated using Compartmental Modeling Software (36,37). To improve the computation efficiency in a high performance computing environment, we rewrote and ran the model using a custom program written in C++ (38) available at (39).

The model state in August 2020, when transmission was steady and low (33,40), was used as a starting point for all scenarios. We introduced a step increase in transmission (transmission hike) on 17 September 2020 to induce a new epidemic wave (Figure 1B). We tested three strengths of the transmission hike (mild, moderate, and strong). Specifically, we hiked the transmission rate parameter from 0.116 to 0.137 (mild), 0.156 (moderate) and 0.179 (strong). The fitted transmission rate in Chicago in its September 2020 wave was between the moderate and strong transmission hikes.

We considered multiple surveillance scenarios for each strength of transmission hike. Each scenario tracked one surveillance indicator: sentinel cases, hospital admissions, or hospital occupancy. Sentinel cases were drawn from incident cases in the mild symptomatic compartment. For sentinel cases and hospital admissions, each indicator was used to calculate the daily instantaneous reproductive number (*R*_*t*_). When *R*_*t*_ exceeded 1.05 for 5 consecutive simulation days, an alarm was triggered. Our preliminary work (See Figure S1 in SI) found that this threshold could trigger mitigations early enough to prevent a transmission surge with relatively low rate of false alarm, i.e., taking mitigative action even before transmission hike, which is seen as inefficient and imprudent. For hospital occupancy, alarm was triggered based on a threshold number of patients in beds. We assume that it would take two days for mitigation to be implemented after an alarm. Once implemented, mitigation immediately reduced the transmission rate to pre-hike levels, and the transmission rate remained the same for the remainder of the simulation period. The drop in transmission to pre-hike level was based on the fitted transmission rates in Chicago after a new round of mitigation policy was imposed to curb the September 2020 wave.

All simulations started 50 days before and ended 150 days after the transmission hike.

### Simulated surveillance and response scenarios

We considered three types of surveillance indicators to guide decision-making: sentinel cases, hospital admissions, and hospital occupancy. For sentinel cases, we considered sampling efforts of 5, 10, 20, 50 and 100%. Sampling effort was defined as the percentage of all mildly symptomatic cases (Sm), regardless of isolation status, that were captured by surveillance. Sentinel cases were calculated by downsampling daily new incident mild symptomatic cases by symptom onset date using a binomial random draw with corresponding probability. Hospital admissions were measured as the daily new incident admissions to the H compartment on each simulation (i.e. the daily rate of flow from compartment SS_i_ to compartment H). Hospital occupancy was the total number of people in the H and Hp compartments.

On each simulation day, the time series of sentinel surveillance indicators from the simulation start to 3 days prior was used to calculate *R*_*t*_ and to evaluate if mitigations should be imposed. The 3-day offset accounted for delays in data collection. For any given symptom onset date, completing the data would require at least 6 days: sentinel cases counted symptomatic people who were tested within 4 days of symptom onset, and we assumed 2 days of test turnaround time. However, this delay can be shortened to 3 days with statistical nowcasting (17,41,42) assuming a 2-day turnaround and that the full numbers of symptomatic cases can be reliably estimated from symptomatic cases within 1 day of symptom onset.

For hospital admissions, we assumed that the data collection delay was 5 days, based on delays associated with admission data in Chicago (17). For hospital occupancy, we assumed data collection delay was only one day. The threshold number of patients in beds for triggering alarm was met when occupancy exceeded 152 per 1 million people for the last 3 days. Occupancy thresholds were chosen based on levels observed in Chicago in October 2020 when mitigative actions were announced.

We estimated *R*_*t*_ using the Python package epyestim (43–45). In this package, the input data series is first bootstrapped and a locally weighted scatterplot smoothing (LOWESS) filter is applied to each of the bootstrapped data series for smoothing. We used a smoothing window of 21 days for all indicators except sentinel surveillance with sampling effort of 5%, which required a 28-day smoothing window to stabilize the *R*_*t*_ trajectory. We approximated the generation time using the serial interval distribution estimated by (35). Time between infection and symptom onset or hospitalization was estimated using Chicago data (see SI for details).

We simulated 500 realizations for each combination of the 3 transmission hike strengths (mild, moderate, and strong); 7 indicators (sentinel cases with sampling effort of 5, 10, 20, 50 and 100%, hospital admission and hospital occupancy); and 3 population sizes (1.25 million, 2.5 million, and 5 million); for a total of 31,500 simulations.

### Timeliness, false alarm rate, deaths averted, and extra days in mitigation

We compared the performance of each indicator by measuring how timely they were in triggering the alarm for taking mitigative actions. Performance was evaluated by calculating the median and 90^th^ percentile of day of triggering alarm among the 500 realizations for each surveillance scenario, as well as the false alarm rate. Lead time of one indicator over another was calculated from the number of days between the median dates (or 90^th^ percentile dates) of triggering alarm. The 90^th^ percentile metric reflected the tail-end performance of the indicators and was less perturbed by false alarms than using median. False alarm rates were the proportion of realizations (out of 500) that met criteria for triggering alarm prior to the date of the transmission hike, hence too early.

To quantify the benefits and costs of using the surveillance indicators in our experiment, we calculated the number of deaths averted and the number of extra days in mitigation in each simulation run. We used hospital occupancy as the reference scenario as it was the slowest to trigger alarm. Each realization in a surveillance scenario was compared against the realization with the same random number seed in the hospital occupancy scenario. We calculated the total number of deaths that occurred throughout the simulation period (50 days before and 150 days after the transmission hike). Deaths averted was the difference between the deaths in the reference and the comparison realizations. The number of extra mitigation days was calculated by subtracting the day of triggering alarm in the comparison realization from that of the reference, which are matched by the random number generator seed. The mean deaths averted and extra mitigation days among all 500 realizations was used to characterize the performance of the surveillance indicator.

We measured the efficiency of each surveillance indicator by calculating the number of deaths averted per extra day of mitigation relative to hospital occupancy. This was calculated by dividing the average deaths averted by average extra mitigation days.

### Age-structured SEIR model

The age-structured model consisted of two SEIR submodels, representing two age groups: below and above 40 years old. The two age groups have similar size in Chicago (55% vs 45%), according to American Community Survey 2016 to 2020 5-year estimates (46). The structure of each submodel was the same as in Figure 1A. We assumed 80% of contacts were within-group and 20% were between-group. This inter-group contact rate is lower than that estimated from pre-pandemic period (47,48) and was chosen to maintain distinctive transmission trends between age groups such that we could evaluate the ability of outpatient sentinel surveillance to detect distinct trends in younger ages. As informed by (23), the probability of asymptomatic infection given exposure was 59% and 19% for the under-40 and above-40 age groups, respectively. The probability a symptomatic infection was severe was 3% and 12% for the under-40 and above-40 age group, respectively, based on CDC’s risk by age group estimates (49). See SI for more details on the parameterization. We assumed that the transmission rate for the above-40 age group was 65% that of the below-40 age group due to a lower overall level of social contact. All other parameters were the same across age groups.

We considered the same scenarios of transmission hikes and surveillance indicators as in the base model. We simulated transmission under two scenarios: one where both age groups experienced the transmission hike simultaneously and one where hike for the above-40 group lagged the hike in the under-40 group by 14 days. We used a population of 2.5 million individuals and a moderate transmission hike. We ran 500 realizations for each surveillance scenario, 2 lags, and 6 surveillance indicators resulting in 500 × 2 × 6 = 6000 simulations. Indicators and criteria for mitigative action were defined as in the base model scenarios. We exclude the hospital occupancy indicator since the purpose of this experiment is to compare the timeliness between sentinel cases and hospital admissions and that no costs and benefits analysis was conducted.

## Results

To compare the performance of sentinel surveillance of outpatient COVID-19 cases compared to hospital-based indicators, we simulated a step increase in transmission (“transmission hike”) and evaluated how quickly and reliably each indicator could trigger an alarm for taking mitigative action while avoiding premature alarms. We compared sentinel surveillance systems that captured between 5% (low sampling effort) and 100% (high sampling effort) of all mild symptomatic cases. For sentinel surveillance and hospital admissions, each data series was used to estimate the instantaneous reproductive number *R*_*t*_, and the threshold for alarm was met when *R*_*t*_ > 1.05 for the previous 5 days. For hospital occupancy, the thresholds were set at exceeding 152 per 1 million for the previous 3 days and were based on levels observed in Chicago in October 2020 when mitigative actions were announced.

### Sentinel surveillance has an operational recency advantage over hospital admissions

Before the transmission hike, all indicators remained steady (Figure 2A). Simulated data series of hospital admissions and sentinel cases with low sampling effort were noisier relative to the mean than for hospital occupancy or sentinel cases with high sampling effort. After the transmission hike, all indicators increased until mitigative action was taken. Sentinel surveillance indicators, which report on newly symptomatic individuals, generally peaked the earliest, followed by hospital admissions and hospital occupancy.

**Figure 2.**
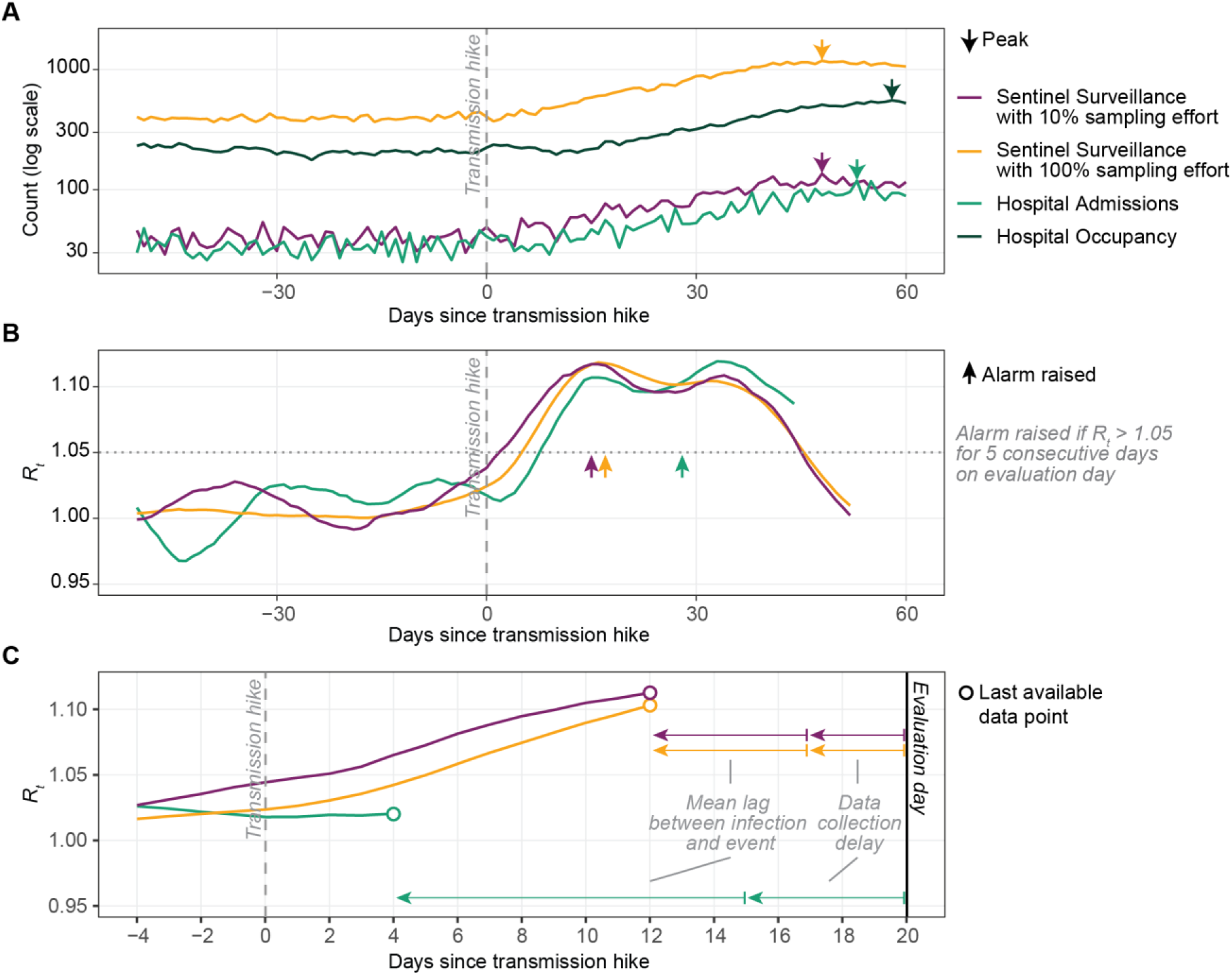
Example trajectories of COVID-19 indicators and their derived *R*_*t*_, from a single, representative simulation with population 2.5 million, moderate transmission hike, and alarm triggered by hospital occupancy. (A) Indicators are steady before and rise rapidly after the transmission hike. Arrows: peak for each indicator. Vertical dashed line: day of transmission hike. (B) *R*_*t*_ estimated based on each indicator, evaluated on day 60 after the transmission hike. Rt could not be estimated using hospital occupancy. Horizontal dotted line: threshold (*R*_*t*_=1.05) for triggering alarm. Arrows: day alarm would have been triggered based on monitoring *R*_*t*_ for each indicator. (C) Real-time evaluation of *R*_*t*_ demonstrates the operational recency advantage of sentinel surveillance over hospital admissions. Example shown for evaluation day of 20 days after the transmission hike.

Data from hospital admissions and from sentinel surveillance with 10% sampling effort produced *R*_*t*_ estimates that fluctuated more than *R*_*t*_ estimates produced from data from sentinel surveillance with 100% sampling effort (Figure 2B). In Figure 2B prior to the transmission hike, *R*_*t*_ estimated from hospital admissions or sentinel surveillance with 10% sampling effort ranged from 0.97 to 1.03 and from 0.99 to 1.04, fluctuating substantially above 1 and below 1 while *R*_*t*_ estimated from sentinel surveillance with 100% sampling effort remained flat at close to 1 (1.0 to 1.02).

On a given evaluation date, *R*_*t*_ cannot be estimated for the most recent 8 days for sentinel surveillance (Figure 2C). For hospital admissions, the most recent 16 days cannot be estimated because the infection-to-hospitalization and the hospitalization-to-report lags are both greater. The modeled 8-day operational recency advantage of sentinel surveillance over hospital admissions is sensitive to the assumed lags and could decrease if data collection delays for hospital data or lag between infection and admission were reduced.

### Sentinel surveillance raises alarms sooner and with a lower false alarm rate

To reduce time spent under mitigation, a good indicator should consistently meet the threshold for triggering alarm closely after, but not before, the transmission hike. False alarms are situations where the threshold was met prior to the transmission hike. For each simulation, we extracted the day on which the criteria for triggering alarm was met.

Compared to hospital admissions or hospital occupancy indicators, use of sentinel surveillance indicators could result in sooner alarms after the transmission hike and a lower rate of false alarms, depending on a few key factors (Figure 3). Sentinel surveillance most outperformed hospitalization indicators for larger changes in transmission, larger populations, or higher sampling effort. In a population of 2.5 million, sentinel surveillance with 10 to 100% sampling effort led hospital admissions in triggering alarms by a median of 2 to 5 days for a mild transmission hike and 6 days for moderate and strong transmission hikes. Although sentinel surveillance with 5% sampling effort led hospital admissions by even more days, this was mainly because of the high false alarm rate. When comparing the 90^th^-percentile day of triggering alarm, sentinel surveillance led hospital admissions by 4 to 7 days for a mild transmission hike and 7 to 9 days for moderate and strong transmission hikes. The lead time of sentinel surveillance with 5 to 100% sampling effort over hospital admissions was similar for populations of 2.5 and 5 million. The lead time advantage in terms of median day of triggering alarm disappeared in the population of 1.25 million, due to higher rates of false alarms. Nevertheless, the lead time advantage in terms of 90^th^-percentile day of triggering alarm remained similar to other population sizes under moderate and strong transmission hikes.

**Figure 3.**
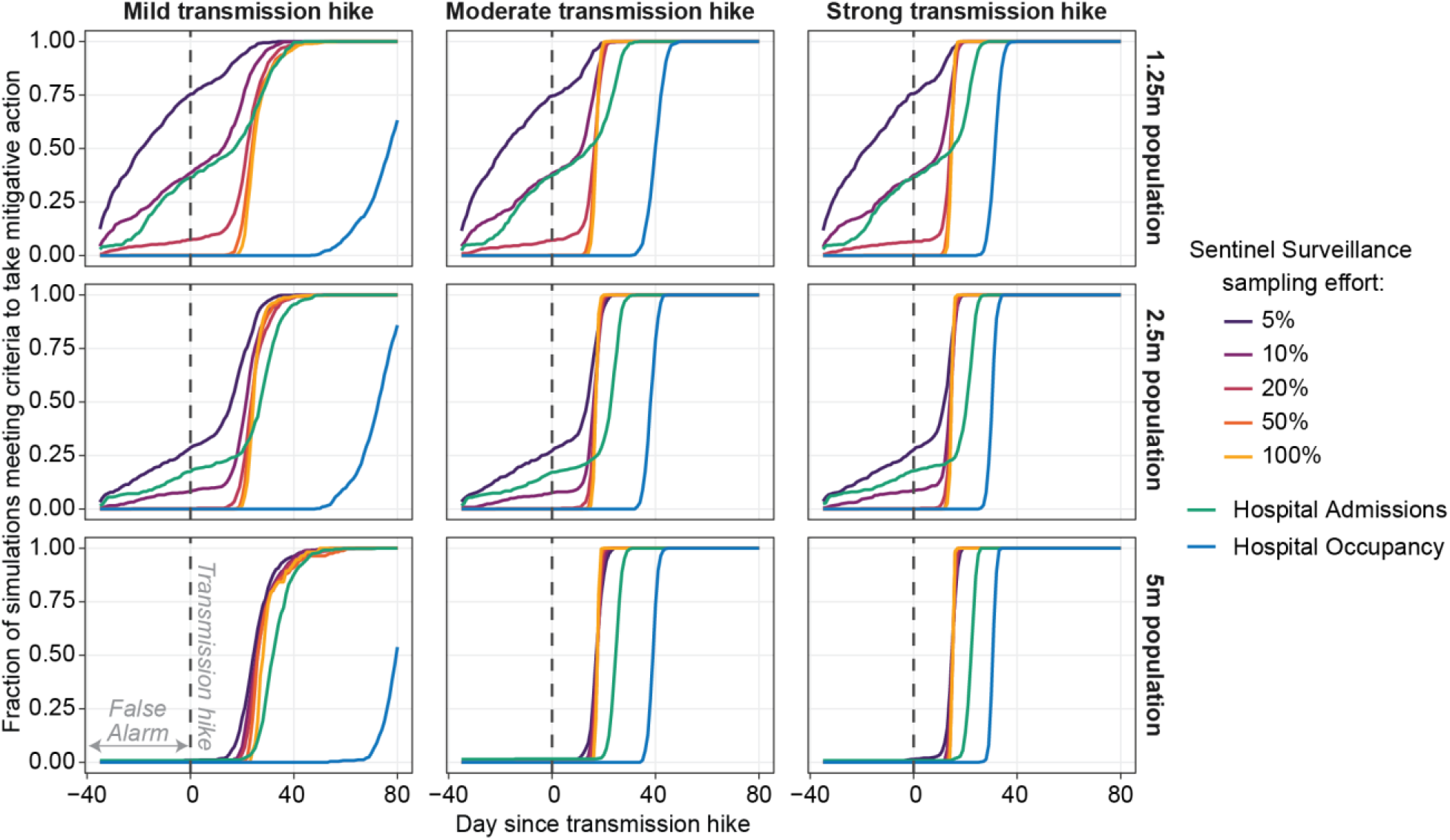
The cumulative distribution of the day on which criteria to trigger alarm was met, by indicator, strengths of transmission hike (columns), and population size (rows).

The false alarm rate, or percentage of realizations that raised a false alarm, was substantial for sentinel surveillance with 5% or 10% sampling effort and for hospital admissions. The false alarm rate was similar regardless of the size of the transmission hike and decreased with increasing sentinel surveillance sampling effort. Across all transmission hikes tested and in a population of 2.5 million, the false alarm rate was 27 to 28%, 8 to 9%, and 17 to 18% respectively for sentinel surveillance sampling effort of 5%, 10%, and for hospital admissions. The false alarm rate was lower in a simulated population of 5 million: sentinel surveillance with 5% sampling effort and hospital admissions each had only 1 to 2% false alarm rate and other indicators had negligible false alarm rates. In a smaller population of 1.25 million, false alarm rates were higher: even sentinel surveillance with 20% sampling effort had a false alarm rate of 7%.

The hospital occupancy indicator resulted in the latest alarm of all indicators considered and did not result in any false alarms. Under the mild transmission hike, alarm was raised 80 days after the transmission hike for 63, 86, and 54% of realizations for populations of 1.25, 2.5 and 5 million respectively.

These results assume that detecting mild symptomatic cases does not lead to isolating behavior and hence lowered infectiousness. We modified the model such that 30% of the mild symptomatic cases were detected and isolated, and that 2/3^rd^ of these detected mild cases were captured by the sentinel surveillance system. We compared the performance of sentinel surveillance with 20% sampling effort and hospital admissions. We found that the outcomes were similar to that of previous results (Figure S2 in SI), suggesting the timeliness advantage of sentinel surveillance persists regardless of isolation.

### Sentinel surveillance of 20% or more effort averts more deaths with fewer extra days of mitigation

To assess the costs and benefits of using sentinel surveillance or hospital admissions indicators to trigger alarms and hence mitigation actions, we compared the number of additional days spent under mitigation (cost) and number of deaths averted (benefit) relative to a baseline scenario with hospital occupancy as indicator. Days under mitigation and deaths were each aggregated over the period from 50 days before to 150 days after the transmission hike. The most efficient indicator would be one that maximizes deaths averted while minimizing extra days of mitigation.

As expected, there was a positive relationship between number of additional days of mitigation and deaths averted (Figure 4A). Spending more time under mitigation always had positive impact on deaths averted. Sentinel surveillance with 20, 50, and 100% sampling effort resulted in similar deaths averted and additional days of mitigation, indicating that increasing sampling effort above 20% did not substantially change system outcomes. Within the same size of transmission hike, decreasing sampling effort for sentinel surveillance, or the use of hospital admission, resulted in larger variation in both deaths averted and additional days of mitigation. The higher variability in using hospital admissions can be attributed to a higher false alarm rate, which imposes mitigation too soon, and late imposition of mitigations.

**Figure 4.**
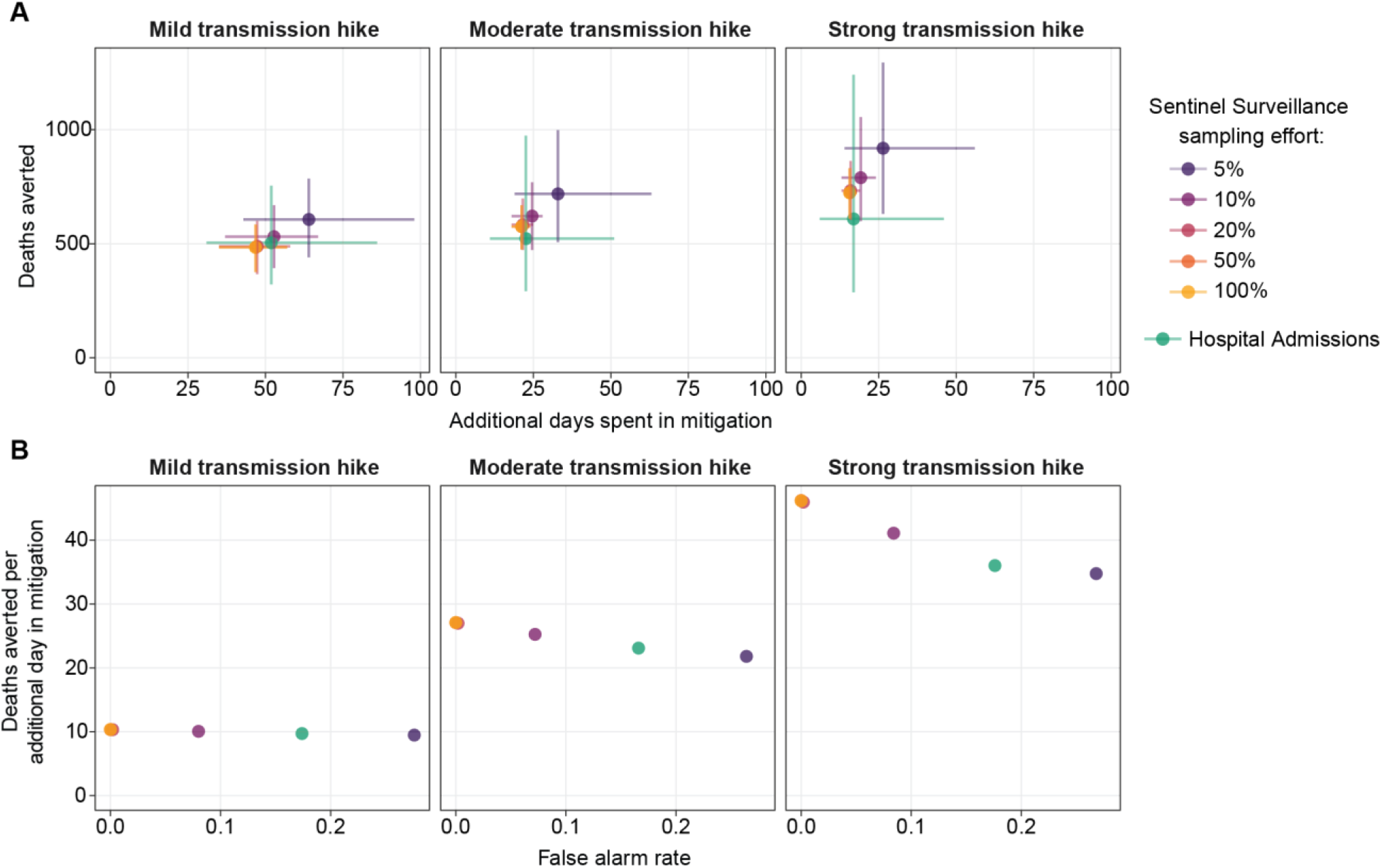
Deaths averted and additional days spent in mitigation for using sentinel surveillance or hospital admissions indicators as the trigger for imposing mitigation measures, under 3 levels of transmission hikes in a population of 2.5 million, compared with using hospital occupancy. (A) Mean and 90% confidence interval of deaths averted and additional days of mitigation, from 500 stochastic realizations per indicator. (B) Average deaths averted per average additional day in mitigation for each indicator and its relationship with false alarm rate. False alarm rate is the proportion of simulations in which action is taken before the transmission hike.

As the size of the transmission hike increased, the number of deaths averted also increased, whereas additional days of mitigation decreased. The mean deaths averted per mean additional day spent in mitigation, which demonstrates the efficiency of mitigation, are shown in Figure 4B. Deaths averted per additional day of mitigation decreased with higher false alarm rates under the moderate and strong transmission hikes. For sentinel surveillance with 20% or more effort, the average deaths averted per additional day of mitigation was 10.3, 26.5 and 46.1 for mild, moderate, and strong transmission hikes respectively. For hospital admissions, the average deaths averted per additional day of mitigation was 9.7, 23.1, and 36.0; for sentinel surveillance with 5% effort average deaths averted per additional day of mitigation was 9.5, 21.8, and 34.8 for mild, moderate, and strong transmission hikes respectively.

### Timeliness advantage of sentinel surveillance widens when transmission increases first in younger ages

Changes in transmission may not affect all age groups simultaneously. We simulated SARS-CoV-2 transmission in an age-structured model that included two age groups, one below and one above 40 years of age. We considered two scenarios: one where both age groups experienced the step increase in transmission simultaneously and one where the step increase for the above-40 group lagged by 14 days (Figure 5A). We compared the performance of sentinel surveillance with various sampling efforts to hospital admissions at responding quickly while minimizing false alarms.

**Figure 5.**
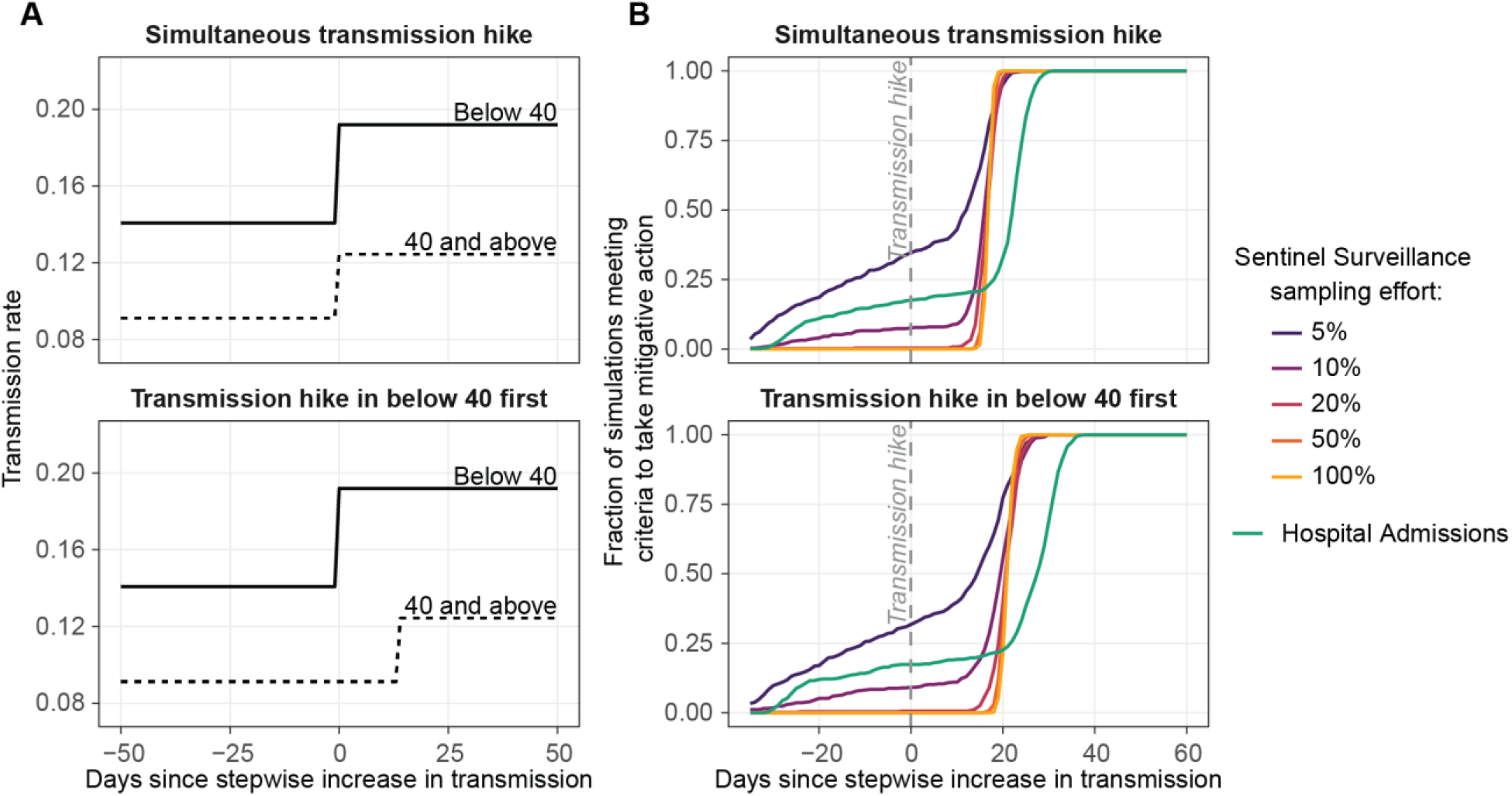
Performance of sentinel surveillance and hospital admissions indicators under scenarios when two age groups (below and above 40 years of age) simultaneously experience a transmission hike or when a transmission hike occurs 14 days earlier in people below 40 years of age. (A) Transmission rate profile for the two scenarios modeled in the age-structured SEIR model. (B) The cumulative distribution of the day on which criteria to trigger alarm was met, using each indicator under the two transmission hike scenarios. Simulations are conducted on population of 2.5 million experiencing a moderate transmission hike with 500 realizations.

Alarm was triggered later in scenarios when the transmission hike occurred first in the under-40 age group (Figure 5B). When the age groups experienced the transmission hike simultaneously, sentinel surveillance with 20% or more effort triggered alarm sooner than hospital admissions by a median of 6 days, and a 90^th^-percentile of 7 days. When the transmission hike occurred first in the under-40 group, sentinel surveillance with greater than 20% sampling led hospital admissions by a median of 7 days, with a 90^th^ percentile lead time of 9 days. Overall, staggering the transmission hike to occur 2 weeks earlier in the younger age group increased the lead time of sentinel surveillance over hospital admissions by 1 to 2 days.

## Discussion

Sentinel surveillance of outpatient recently-symptomatic cases provides a useful indicator to alert decision-makers about increases in SARS-CoV-2 transmission. Compared with hospital admissions, sentinel surveillance is less variable, reduces the false alarm rate, and provides data with shorter lag times, which allows faster reactions to changes. The minimal sampling effort required for reliable sentinel surveillance indicators varies according to the population size and size of transmission increase, but generally capturing 20% of mild symptomatic cases within the surveillance system was sufficient.

The 8-day operational recency of outpatient sentinel surveillance over hospital admissions—that sentinel surveillance can provide up to 8 days’ advance warning of changes in transmission compared with hospital admissions data—can be attributed to shorter delay between infection and symptom onset, as well as the use of statistical nowcasting. Nowcasting has been widely used to estimate the complete case count using incomplete time series for COVID-19 and other diseases (41,42,50–52). In Chicago, nowcasting could be applied to shorten the data collection delay for sentinel cases, but not hospital admissions due to inconsistent backfilling (17). Improvements in reporting speed of hospital admissions data would reduce the operational recency of sentinel surveillance to as little as 5 days.

Our study highlights that indicators with small counts, e.g., less than 50 per day, can have limited utility in decision making due to their high variability, which leads to a high false alarm rate. These factors can result in unnecessary restrictions, reduce decision makers’ confidence in taking action, and harm public trust in the health system. At a given population size, small counts in sentinel surveillance can be overcome by increasing sampling effort. On the other hand, hospital admissions are inherently limited by severe disease rates of COVID-19.

We defined the threshold for triggering alarm as *R*_*t*_ > 1.05 for 5 consecutive days. Setting the threshold higher would have reduced the number of deaths averted and decreased the number of days spent in mitigation, whereas a lower threshold would have led to more false alarms. 1.05 was selected as an acceptable middle ground. We expect that factors affecting sentinel surveillance performance would be similar if a different threshold were chosen.

While false alarms may incur economic costs or reduce public confidence in the decision-making process, they resulted in the most deaths averted. However, false alarms reduced the efficiency of mitigative action, measured as deaths averted per additional day spent in mitigation. When we removed simulations with false alarms, efficiency of surveillance based on hospital admissions increased or slightly surpassed the efficiency of sentinel surveillance (Figure S3 in SI). Nevertheless, the overall deaths averted for hospital admissions was 25 to 35% lower than for high-effort sentinel surveillance under a moderate or strong increase in transmission. At similar efficiency, the operational recency of sentinel surveillance over hospital admissions led to saving more lives.

Hospitalization indicators are more effective at monitoring the transmission in older age groups than in the population at-large. For example, 80% of all COVID-19 hospitalizations in Chicago were older than 40 years old whereas only 50% of the general population was older than 40 years (53). Outpatient sentinel surveillance thus holds an additional timeliness advantage over hospital admissions when the younger population experiences an increase in transmission earlier than the older population. Situations like this were common throughout this pandemic, e.g., in Illinois and Florida, USA, during their winter and summer 2020 wave (18), and in England UK during the Omicron wave in late 2021 (19). Sentinel surveillance for early warning could be especially useful when transmission first increases in demographic group less prevalent in hospitalization data.

We chose a between-age group contact rate of 20% to ensure that cases and hospitalizations in those age below 40 were distinctively elevated before those above 40. More contacts between age groups would disintegrate this pattern and erase the additional advantage conveyed by the sentinel surveillance over hospital admissions; fewer contacts would boost the advantage. This between-group contact rate is lower than the estimate of 30% that is inferred from pre-pandemic age contact matrices (47,48), which might have changed significantly due to the impact of public health measures and reduction of number of social contacts (54).

A major challenge in implementing sentinel surveillance is to achieve the minimal sampling effort needed for decision making. In USA and other countries where tests were increasingly abundant after the early weeks of pandemic, attaining 20% sampling effort could have been feasible provided that all outpatient testing sites were reliably recording symptom status and onset dates. The ascertainment rate for symptomatic illnesses exceeded 30% in these countries (55–59). Since at-home antigen testing has become widely available in many locales, new approaches to surveillance will likely be needed. For example, population-based survey of symptom information, such as that of UK’s Office of National Statistics (60), could be implemented to aid this sentinel surveillance model. Self-reported symptom tracking programs such as ZOE health study (61), COVID Symptom (62) and COVID Control (63) may compliment the surveillance model, although maintaining their longevity and public interests remain challenging (64).

If questions on patients’ symptom status and symptom onset date were embedded into patient intake forms since the inception of the testing programs, then the cost of implementing outpatient sentinel surveillance would be very low. However, changing an existing system to incorporate new questions may incur significant technological and coordination costs, as it would require a change in frontend surveys and databases across multiple testing vendors.

Our study did not compare the utility of sentinel surveillance against reported cases and test positivity rates, despite the latter two indicators’ prominent role in the COVID-19 pandemic. Simulating the time-varying biases in cases and test positivity rate is challenging due to the lack of understanding in these indicators’ biases over time, and demand for tests can fluctuate in response to population perception of transmission rates. Implementing outpatient sentinel surveillance can provide important data to correct the biases in cases and test positivity rates, increasing their utility for decision-making.

## Conclusions

Our study shows that sentinel surveillance of recently-symptomatic cases in outpatient testing sites could be feasible, prudent, and effective for informing situational awareness in an epidemic like COVID-19. With adequate sampling effort of at least 20% of mild symptomatic cases captured by surveillance, sentinel cases provide accurate, timely warning of increases in transmission, even under heterogeneous transmission conditions. In practice, decisions should not be based on only one indicator: cross-checking with other indicators will provide more confidence that action should or should not be taken.

## Data Availability

All data produced in the present work are contained in the manuscript.

## List of abbreviations

USA: United States of America
UK: United Kingdom
SEIR: Susceptible exposed infected recovered
ICU: Intensive care unit
LOWESS: Locally weighted scatterplot smoothing
CDC: Centers for Disease Control and Prevention

## Declarations

### Ethics approval and consent to participate

Not applicable

### Consent for publication

Not applicable

### Availability of data and materials

Code used to simulate SARS-CoV-2 transmission and sentinel surveillance are available at *https://github.com/numalariamodeling/covid-sentinel-surveil*. Datasets used in this study were generated using the code in the repository.

### Authors’ contributions

Conceived the study: KBT, MR, JG. Developed model and code: KBT, MR, RR, TH. Wrote main manuscript text: KBT, JG. All authors edited and reviewed the manuscript.

## Acknowledgments

We thank Jiayi Gu, Josephine Harter, Vikram Thanigaivelan, Laith Kassiseh and Jamie Woodworth for their preliminary work. This study would not have been possible without the computational resources and staff support from the Quest high performance computing facility at Northwestern University.

## Supplementary Information

### Additional file 1 — Details of SEIR model fitting and parameters, and supplemental figures

Specific values for the model parameters, and figures showing (a) timeliness performance of sentinel surveillance and hospital admissions when detection of mild cases leads to isolation, and (b) efficiency of surveillance indicators when stochastic realizations with false alarms for hospital admissions were removed.

## References

1. WHO. WHO Coronavirus (COVID-19) Dashboard [Internet]. [cited 2022 Aug 16]. Available from: https://covid19.who.int

2. Dong E, Du H, Gardner L. An interactive web-based dashboard to track COVID-19 in real time. The Lancet Infectious Diseases. 2020 May;20(5):533–4.

3. Ahlers M, Aralis H, Tang W, Sussman JB, Fonarow GC, Ziaeian B. Non-pharmaceutical interventions and covid-19 burden in the United States: retrospective, observational cohort study. BMJ Medicine. 2022 Aug 1;1:e000030.

4. Leech G, Rogers-Smith C, Monrad JT, Sandbrink JB, Snodin B, Zinkov R, et al. Mask wearing in community settings reduces SARS-CoV-2 transmission. Proc Natl Acad Sci USA. 2022 Jun 7;119(23):e2119266119.

5. Liu X, Xu X, Li G, Xu X, Sun Y, Wang F, et al. Differential impact of non-pharmaceutical public health interventions on COVID-19 epidemics in the United States. BMC Public Health. 2021 May 21;21(1):965.

6. Rebmann T, Loux TM, Arnold LD, Charney R, Horton D, Gomel A. SARS-CoV-2 Transmission to Masked and Unmasked Close Contacts of University Students with COVID-19 — St. Louis, Missouri, January-May 2021. MMWR Morb Mortal Wkly Rep. 2021 Sep 10;70(36):1245–8.

7. Van Dyke ME, Rogers TM, Pevzner E, Satterwhite CL, Shah HB, Beckman WJ, et al. Trends in County-Level COVID-19 Incidence in Counties With and Without a Mask Mandate — Kansas, June 1–August 23, 2020. MMWR Morb Mortal Wkly Rep. 2020 Nov 27;69(47):1777–81.

8. Yang H, Sürer Ö, Duque D, Morton DP, Singh B, Fox SJ, et al. Design of COVID-19 staged alert systems to ensure healthcare capacity with minimal closures. Nature Communications 2021 12:1. 2021 Jun;12(1):1–7.

9. Ibrahim NK. Epidemiologic surveillance for controlling Covid-19 pandemic: types, challenges and implications. J Infect Public Health. 2020 Nov;13(11):1630–8.

10. Rader B, Astley CM, Sy KTL, Sewalk K, Hswen Y, Brownstein JS, et al. Geographic access to United States SARS-CoV-2 testing sites highlights healthcare disparities and may bias transmission estimates. J Travel Med. 2020 Nov 9;27(7):taaa076.

11. Moss R, Zarebski AE, Carlson SJ, McCaw JM. Accounting for Healthcare-Seeking Behaviours and Testing Practices in Real-Time Influenza Forecasts. Trop Med Infect Dis. 2019 Jan 11;4(1):E12.

12. Tian S, Chang Z, Wang Y, Wu M, Zhang W, Zhou G, et al. Clinical Characteristics and Reasons for Differences in Duration From Symptom Onset to Release From Quarantine Among Patients With COVID-19 in Liaocheng, China. Front Med. 2020;7:210.

13. Alene M, Yismaw L, Assemie MA, Ketema DB, Gietaneh W, Birhan TY. Serial interval and incubation period of COVID-19: a systematic review and meta-analysis. BMC Infectious Diseases. 2021 Mar 11;21(1):257.

14. Lauer SA, Grantz KH, Bi Q, Jones FK, Zheng Q, Meredith HR, et al. The Incubation Period of Coronavirus Disease 2019 (COVID-19) From Publicly Reported Confirmed Cases: Estimation and Application. Ann Intern Med. 2020 May 5;172(9):577–82.

15. Yang X, Yu Y, Xu J, Shu H, Xia J, Liu H, et al. Clinical course and outcomes of critically ill patients with SARS-CoV-2 pneumonia in Wuhan, China: a single-centered, retrospective, observational study. The Lancet Respiratory Medicine. 2020 May 1;8(5):475–81.

16. Gostic KM, McGough L, Baskerville EB, Abbott S, Joshi K, Tedijanto C, et al. Practical considerations for measuring the effective reproductive number, Rt. PLOS Computational Biology. 2020 Dec 10;16(12):e1008409.

17. Richardson R, Jorgensen E, Arevalo P, Holden TM, Gostic KM, Pacilli M, et al. Tracking changes in SARS-CoV-2 transmission with a novel outpatient sentinel surveillance system in Chicago, USA. Nat Commun. 2022 Sep 22;13(1):5547.

18. Holden TM, Richardson RAK, Arevalo P, Duffus WA, Runge M, Whitney E, et al. Geographic and demographic heterogeneity of SARS-CoV-2 diagnostic testing in Illinois, USA, March to December 2020. BMC Public Health. 2021 Dec;21(1):1–13.

19. Office for National Statistics. Coronavirus (COVID-19) Infection Survey, UK: 14 January 2022 [Internet]. [cited 2022 Aug 16]. Available from: https://www.ons.gov.uk/peoplepopulationandcommunity/healthandsocialcare/conditionsanddiseases/bulletins/coronaviruscovid19infectionsurveypilot/14january2022#age-analysis-of-the-number-of-people-who-had-covid-19

20. Campillo-Funollet E, Van Yperen J, Allman P, Bell M, Beresford W, Clay J, et al. Predicting and forecasting the impact of local outbreaks of COVID-19: use of SEIR-D quantitative epidemiological modelling for healthcare demand and capacity. Int J Epidemiol. 2021 Aug 30;50(4):1103–13.

21. Delli Compagni R, Cheng Z, Russo S, Van Boeckel TP. A hybrid Neural Network-SEIR model for forecasting intensive care occupancy in Switzerland during COVID-19 epidemics. PLOS One. 2022;17(3):e0263789.

22. Moghadas SM, Shoukat A, Fitzpatrick MC, Wells CR, Sah P, Pandey A, et al. Projecting hospital utilization during the COVID-19 outbreaks in the United States. Proceedings of the National Academy of Sciences. 2020 Apr 21;117(16):9122–6.

23. Davies NG, Klepac P, Liu Y, Prem K, Jit M, Eggo RM. Age-dependent effects in the transmission and control of COVID-19 epidemics. Nat Med. 2020 Aug;26(8):1205–11.

24. Reiner RC, Barber RM, Collins JK, Zheng P, Adolph C, Albright J, et al. Modeling COVID-19 scenarios for the United States. Nat Med. 2021 Jan;27(1):94–105.

25. Qiu T, Xiao H, Brusic V. Estimating the Effects of Public Health Measures by SEIR(MH) Model of COVID-19 Epidemic in Local Geographic Areas. Front Public Health. 2021;9:728525.

26. Berger D, Herkenhoff K, Huang C, Mongey S. Testing and reopening in an SEIR model. Review of Economic Dynamics. 2022 Jan 1;43:1–21.

27. Chang S, Pierson E, Koh PW, Gerardin J, Redbird B, Grusky D, et al. Mobility network models of COVID-19 explain inequities and inform reopening. Nature. 2021 Jan;589(7840):82–7.

28. Rawson T, Brewer T, Veltcheva D, Huntingford C, Bonsall MB. How and When to End the COVID-19 Lockdown: An Optimization Approach. Frontiers in Public Health. 2020;8:262.

29. Abernethy GM, Glass DH. Optimal COVID-19 lockdown strategies in an age-structured SEIR model of Northern Ireland. Journal of The Royal Society Interface. 19(188):20210896.

30. Smith DRM, Duval A, Pouwels KB, Guillemot D, Fernandes J, Huynh BT, et al. Optimizing COVID-19 surveillance in long-term care facilities: a modelling study. BMC Med. 2020 Dec 8;18(1):386.

31. Larremore DB, Wilder B, Lester E, Shehata S, Burke JM, Hay JA, et al. Test sensitivity is secondary to frequency and turnaround time for COVID-19 screening. Science Advances. 2021 Jan;7(1):eabd5393.

32. Lokuge K, Banks E, Davis S, Roberts L, Street T, O’Donovan D, et al. Exit strategies: optimising feasible surveillance for detection, elimination, and ongoing prevention of COVID-19 community transmission. BMC Medicine. 2021 Feb 17;19(1):50.

33. Runge M, Richardson RAK, Clay PA, Bell A, Holdenid TM, Singam M, et al. Modeling robust COVID-19 intensive care unit occupancy thresholds for imposing mitigation to prevent exceeding capacities. PLOS Global Public Health. 2022 May;2(5):e0000308.

34. Wang D, Hu B, Hu C, Zhu F, Liu X, Zhang J, et al. Clinical Characteristics of 138 Hospitalized Patients With 2019 Novel Coronavirus-Infected Pneumonia in Wuhan, China. JAMA. 2020 Mar 17;323(11):1061–9.

35. Bi Q, Wu Y, Mei S, Ye C, Zou X, Zhang Z, et al. Epidemiology and transmission of COVID-19 in 391 cases and 1286 of their close contacts in Shenzhen, China: a retrospective cohort study. The Lancet Infectious Diseases. 2020 Aug;20(8):911–9.

36. The Institute for Disease Modeling. Compartmental Modeling Software (CMS) [Internet]. 2018 [cited 2022 Aug 25]. Available from: https://docs.idmod.org/projects/cms/en/latest/index.html#

37. Northwestern University Malaria Modelling Team. Modelling the COVID-19 pandemic in Illinois [Internet]. NU Malaria Modeling Team; 2021 [cited 2022 Aug 25]. Available from: https://github.com/numalariamodeling/covid-chicago

38. Hladish T, Melamud E, Barrera LA, Galvani A, Meyers LA. EpiFire: An open source C++ library and application for contact network epidemiology. BMC Bioinformatics. 2012 May 4;13(1):76.

39. Northwestern University Malaria Modelling Team. Modeling sentinel surveillance [Internet]. 2021 [cited 2022 Aug 25]. Available from: https://github.com/numalariamodeling/covid-sentinel-surveil

40. City of Chicago. Latest Data | COVID 19 [Internet]. [cited 2022 Aug 17]. Available from: https://www.chicago.gov/city/en/sites/covid-19/home/covid-dashboard.html

41. McGough SF, Johansson MA, Lipsitch M, Menzies NA. Nowcasting by Bayesian Smoothing: A flexible, generalizable model for real-time epidemic tracking. PLOS Computational Biology. 2020 Apr;16(4):e1007735.

42. Höhle M, an der Heiden M. Bayesian nowcasting during the STEC O104:H4 outbreak in Germany, 2011. Biometrics. 2014;70(4):993–1002.

43. Hilfiker L, Josi J. epyestim [Internet]. 2020. Available from: https://pypi.org/project/epyestim/

44. Huisman JS, Scire J, Angst DC, Li J, Neher RA, Maathuis MH, et al. Estimation and worldwide monitoring of the effective reproductive number of SARS-CoV-2. eLife. 2022 Aug;11:e71345.

45. Cori A, Ferguson NM, Fraser C, Cauchemez S. A new framework and software to estimate time-varying reproduction numbers during epidemics. Am J Epidemiol. 2013 Nov 1;178(9):1505–12.

46. United States Census Bureau. Census Bureau Data [Internet]. [cited 2022 Aug 16]. Available from: https://data.census.gov/cedsci/

47. Prem K, Cook AR, Jit M. Projecting social contact matrices in 152 countries using contact surveys and demographic data. PLOS Computational Biology. 2017 Sep 12;13(9):e1005697.

48. Mistry D, Litvinova M, Pastore y Piontti A, Chinazzi M, Fumanelli L, Gomes MFC, et al. Inferring high-resolution human mixing patterns for disease modeling. Nat Commun. 2021 Jan 12;12(1):323.

49. CDC. Risk for COVID-19 Infection, Hospitalization, and Death By Age Group [Internet]. [cited 2022 Aug 18]. Available from: https://www.cdc.gov/coronavirus/2019-ncov/covid-data/investigations-discovery/hospitalization-death-by-age.html

50. Li T, White LF. Bayesian back-calculation and nowcasting for line list data during the COVID-19 pandemic. PLOS Computational Biology. 2021 Jul 12;17(7):e1009210.

51. Bastos LS, Economou T, Gomes MFC, Villela DAM, Coelho FC, Cruz OG, et al. A modelling approach for correcting reporting delays in disease surveillance data. Statistics in Medicine. 2019 Sep;38(22):4363–77.

52. van de Kassteele J, Eilers PHC, Wallinga J. Nowcasting the Number of New Symptomatic Cases During Infectious Disease Outbreaks Using Constrained P-spline Smoothing. Epidemiology. 2019 Sep;30(5):737–45.

53. City of Chicago. Daily COVID-19 Hospitalizations by Age | Chicago Data Portal [Internet]. [cited 2022 Aug 17]. Available from: https://data.cityofchicago.org/Health-Human-Services/Daily-COVID-19-Hospitalizations-by-Age/g43c-xce5

54. Drolet M, Godbout A, Mondor M, Béraud G, Drolet-Roy L, Lemieux-Mellouki P, et al. Time trends in social contacts before and during the COVID-19 pandemic: the CONNECT study. BMC Public Health. 2022 May 23;22:1032.

55. CDC. Estimated COVID-19 Burden [Internet]. [cited 2022 Aug 16]. Available from: https://www.cdc.gov/coronavirus/2019-ncov/cases-updates/burden.html

56. Reese H, Iuliano AD, Patel NN, Garg S, Kim L, Silk BJ, et al. Estimated Incidence of Coronavirus Disease 2019 (COVID-19) Illness and Hospitalization—United States, February– September 2020. Clinical Infectious Diseases. 2021 Jun 15;72(12):e1010–7.

57. Iuliano AD, Chang HH, Patel NN, Threlkel R, Kniss K, Reich J, et al. Estimating under-recognized COVID-19 deaths, United States, march 2020-may 2021 using an excess mortality modelling approach. The Lancet Regional Health – Americas. 2021 Sep 1;1:100019.

58. Pullano G, Di Domenico L, Sabbatini CE, Valdano E, Turbelin C, Debin M, et al. Underdetection of cases of COVID-19 in France threatens epidemic control. Nature. 2021 Feb;590(7844):134–9.

59. Omori R, Mizumoto K, Nishiura H. Ascertainment rate of novel coronavirus disease (COVID-19) in Japan. International Journal of Infectious Diseases. 2020 Jul 1;96:673–5.

60. Pouwels KB, House T, Pritchard E, Robotham JV, Birrell PJ, Gelman A, et al. Community prevalence of SARS-CoV-2 in England from April to November, 2020: results from the ONS Coronavirus Infection Survey. The Lancet Public Health. 2021 Jan 1;6(1):e30–8.

61. ZOE. ZOE Health Study [Internet]. [cited 2022 Aug 18]. Available from: https://health-study.joinzoe.com/

62. COVID Symptom. COVID Symptom [Internet]. [cited 2022 Aug 18]. Available from: https://www.covidsymptom.org/

63. COVID Control. COVID Control-A Johns Hopkins University Study [Internet]. [cited 2022 Aug 18]. Available from: https://covidcontrol.jhu.edu/

64. Janvrin ML, Korona-Bailey J, Koehlmoos TP. Re-examining COVID-19 Self-Reported Symptom Tracking Programs in the United States: Updated Framework Synthesis. JMIR Form Res. 2021 Dec 6;5(12):e31271.

