## Supplemental Information for "Design of effective outpatient sentinel surveillance for COVID-19 decision-making: a modeling study"

### SEIR model fitting and parameters

For most of the parameters required in this model, the suitable range of their values were identified from published figures in the literature and modelers assumptions. Then the parameters were fit to the reported deaths, ICU and non-ICU hospitalizations, and hospital admissions in the city of Chicago, USA. For this study, we took only the portion from 2020 Feb 13 to 2020 July 29 of previous work (1). The fitting was done by running the model repeatedly using different combination of values sampled from the initial range identified. Data for fitting were obtained from the Illinois Department of Public Health.

For the fitting purpose, some parameters were allowed to vary over time via stepwise changes: transmissibility, detection rate of asymptomatic, presymptomatic, mild and severe symptomatic, fraction of people developing critical illness, fatality rate and time to recovery from critical illness. The change points for these parameters are determined based on data in Chicago, or on regular interval (e.g., twice per month for transmission rates). More information about the model fitting and calibration process can be found in (1).

Table S1 details the fitted or assumed values for the model parameters. These values are then used for all simulations in this study, which used the model state in 2020 July 30 as starting point. From this point onwards, we assume the same values for all time varying parameters (using the "final" values in Table S1) except for the transmission rate. We altered the transmission rate as described in the main text to produce different levels of transmission hike.

**Table S1.** Estimates used to simulate SARS-CoV-2 transmission from 2020 Feb 13 to 2020 July 29 after model fitting. All values were fitted unless specified in the remark.

| Parameter | Estimate | Remark |
|---|---|---|
| Fraction symptomatic | 0.59 | (2) |
| Fraction of asymptomatic | 0.41 | Compliment of fraction symptomatic |
| Probability of severe symptom given symptomatic | 0.08 | (3) |
| Probability of mild symptom given symptomatic | 0.92 | Compliment of severe probability |
| Detection rate of mild symptomatic infections (time varying) | Initial: 0.001<br>Final: 0.15 | Calculated from line list data as described in (4) |

| | | |
|---|---|---|
| Detection rate of severe infections (time varying) | Initial: 0.01<br>Final: 0.55 | Calculated from Line List data as described in (4) |
| Probability of critically ill given severe (time varying) | Initial: 0.49<br>Final: 0.15 | (5) |
| Probability of death given critical (time varying) | Initial: 0.20<br>Final: 0.18 | Calculated from case fatality ratio in (6) and probability of critical. |
| Time exposed to infectious (either pre-symptomatic or asymptomatic) | 3.7 days | (7) |
| Time pre-symptomatic to mild or severe symptoms | 3.4 days | (7,8) |
| Time to detection and isolation for severe symptomatic | 2 | Assumed |
| Time to hospitalization from severe symptoms | 4.1 days | (9) |
| Time to critical from hospitalization | 5.6 days | (10) |
| Time to deaths from critical | 5.5 days | (11) |
| Recovery rate of asymptomatic infections | 9 days | Assumed to be the same as mild symptomatic |
| Recovery rate mild symptomatic infections | 9 days | (12) |
| Recovery rate of hospitalized but not critical cases (time varying) | Initial: 5.8 days<br>Final: 4.7 days | (9) |
| Recovery rate of critical cases | 9.7 days | (12) |
| Reduced infectiousness when isolated | 0.01 | Fitted |
| Reduced infectiousness when asymptomatic | 0.8 | Assumed |
| Transmission rate (from susceptible to exposed) | Initial: 1.05<br>Final: 0.12 | Fitted |

### Supplemental results and discussions figures

*Effects of triggering criteria on timeliness in raising the alarm of each indicator*

For sentinel surveillance and hospital admissions indicators, we chose to trigger the alarm (for imposing mitigations) when the derived Rt exceeded 1.05 for 5 consecutive simulation days. This is based on our preliminary work that the threshold has relatively low rate of false alarm, and yet could trigger mitigations early enough to prevent a transmission surge.

In Fig. S1, we compare the quality of raising alarm using three different criteria, Rt exceeding 1.05 for 3, 5 and 7 consecutive days. False alarm rate reduces, and the median day of triggering actions increases, when more consecutive days are needed. We found both 5 and 7 consecutive days acceptable.

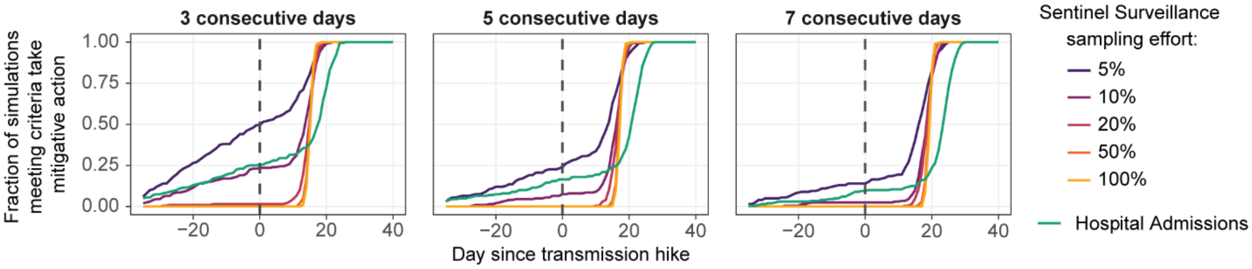


**Figure S1**. The cumulative distribution of the day of triggering alarm, when the threshold was set to 3, 5 or 7 consecutive days of indicators derived Rt exceeding 1.05. Simulations are conducted on a population of 2.5 million experiencing a moderate transmission hike, with 200 stochastic realizations per indicator.

*Timeliness advantage of sentinel surveillance persists when detection of mild cases leads to isolation*

Results in the main text assumed that detecting mild symptomatic cases does not lead to isolating behavior and hence lowered infectiousness. However, people who test positive for SARS-CoV-2 are probably more likely to isolate, and thus a sentinel surveillance system with greater sampling effort may reduce transmission just by detecting more infections. This may affect the overall timeliness results.

We modified the model such that the detection probability for mildly symptomatic cases was fixed at 30% and detection probability of asymptomatic and presymptomatic infections was fixed at 5%. We assumed the sentinel surveillance system could achieve a maximum of 20% sampling effort, which would correspond to the case where 2/3rd of the detected mildly symptomatic cases were captured in the surveillance system.

In the model that included isolation, the timeliness advantage of sentinel surveillance with 20% sampling effort over hospital admissions remained unchanged. The median lead time was 6 days, and the 90$^{th}$-percentile was 9 days, similar to previous simulations where detected mildly symptomatic cases did not isolate (Figure S2).

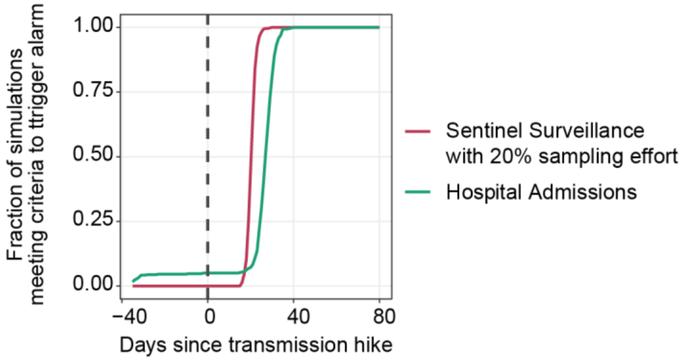


**Figure S2**. The cumulative distribution of the day on which criteria to trigger alarm was met, using sentinel surveillance with 20% sampling effort or hospital admissions as indicator, if detected cases isolate. Simulations are conducted on a population of 2.5 million experiencing a moderate transmission hike, with 500 stochastic realizations per indicator.

*Efficiency of hospital admissions increases when stochastic realizations with false alarms are removed*

As a whole, we showed that hospital admissions averted less deaths per extra mitigation day. This is attributable to false alarms. The efficiency of hospital admissions is similar or slightly higher than sentinel surveillance if we remove the stochastic realizations with false alarms for hospital admissions (Figure S3). However, hospital admissions with similar efficiency as sentinel surveillance averted substantially less overall deaths, highlighting the disadvantage of additional operational latency.

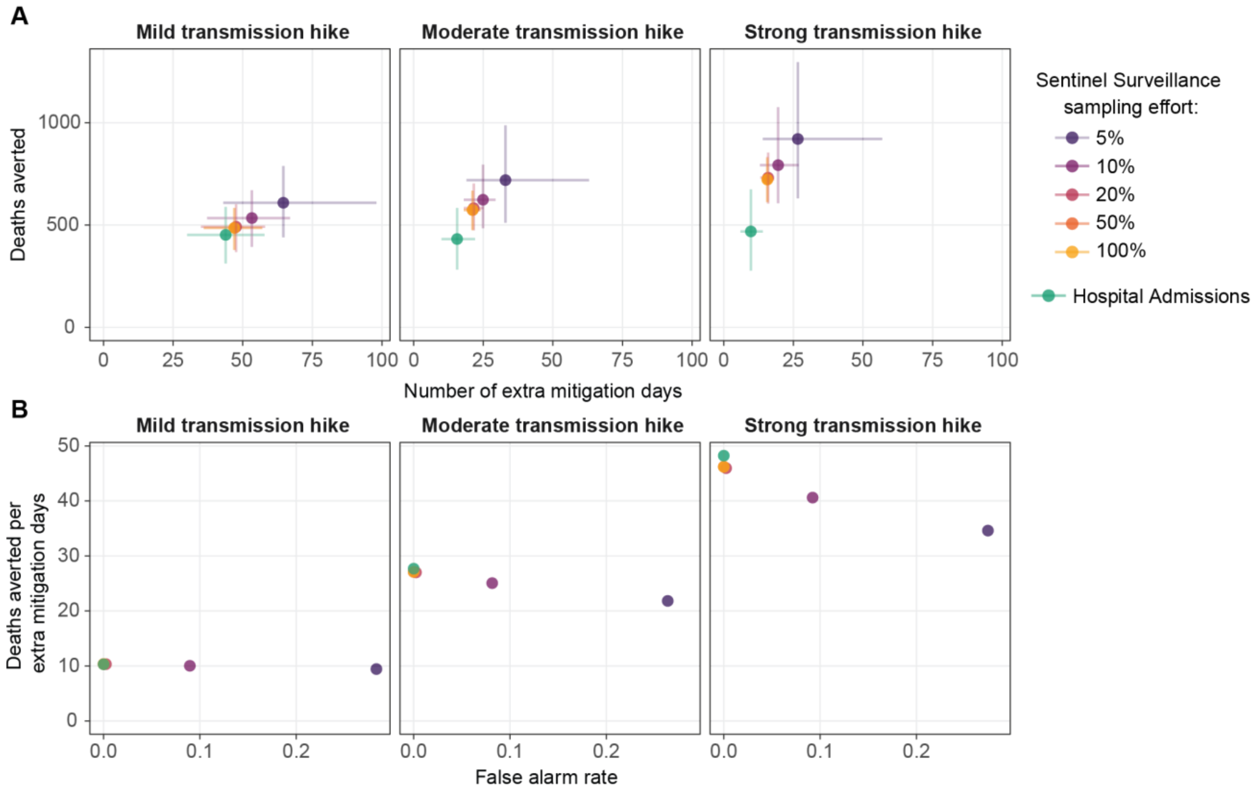


**Figure S3.** Deaths averted and additional days spent in mitigation for using sentinel surveillance or hospital admissions indicators as the trigger for imposing mitigation measures, under three wave strengths in a population of 2.5 million, compared with using hospital occupancy. In contrast to Figure 4, stochastic realizations with false alarms when using hospital admissions (~80 out of 500) were excluded from this analysis. (A) Mean and 90% confidence interval of deaths averted and additional days of mitigation, from ~420 stochastic realizations per indicator. (B) Average deaths averted per average additional day in mitigation for each indicator and its relationship with false alarm rate. False alarm rate is the proportion of simulations in which action is taken before the transmission hike.

### References


1. Runge M, Richardson RAK, Clay PA, Bell A, Holdenid TM, Singam M, et al. Modeling robust COVID-19 intensive care unit occupancy thresholds for imposing mitigation to prevent exceeding capacities. PLOS Global Public Health. 2022 May;2(5):e0000308.

2. Oran DP, Topol EJ. Getting a handle on asymptomatic SARS-CoV-2 infection [Internet]. 2020 [cited 2022 Aug 26]. Available from: https://www.scripps.edu/science-and-medicine/translational-institute/about/news/sarc-cov-2-infection/index.html

3. Salje H, Tran Kiem C, Lefrancq N, Courtejoie N, Bosetti P, Paireau J, et al. Estimating the burden of SARS-CoV-2 in France. Science. 2020 Jul 10;369(6500):208–11.

4. Holden TM, Richardson RAK, Arevalo P, Duffus WA, Runge M, Whitney E, et al. Geographic and demographic heterogeneity of SARS-CoV-2 diagnostic testing in Illinois, USA, March to December 2020. BMC Public Health. 2021 Dec;21(1):1–13.

5. Lewnard JA, Liu VX, Jackson ML, Schmidt MA, Jewell BL, Flores JP, et al. Incidence, clinical outcomes, and transmission dynamics of severe coronavirus disease 2019 in California and Washington: prospective cohort study. BMJ. 2020 May 22;369:m1923.

6. Oke J, Heneghan C. Global Covid-19 Case Fatality Rates [Internet]. The Centre for Evidence-Based Medicine. [cited 2022 Aug 26]. Available from: https://www.cebm.net/covid-19/global-covid-19-case-fatality-rates/

7. Li R, Pei S, Chen B, Song Y, Zhang T, Yang W, et al. Substantial undocumented infection facilitates the rapid dissemination of novel coronavirus (SARS-CoV-2). Science. 2020 May 1;368(6490):489–93.

8. Qin J, You C, Lin Q, Hu T, Yu S, Zhou XH. Estimation of incubation period distribution of COVID-19 using disease onset forward time: A novel cross-sectional and forward follow-up study. Sci Adv. 2020 Aug;6(33):eabc1202.

9. Wang D, Hu B, Hu C, Zhu F, Liu X, Zhang J, et al. Clinical Characteristics of 138 Hospitalized Patients With 2019 Novel Coronavirus-Infected Pneumonia in Wuhan, China. JAMA. 2020 Mar 17;323(11):1061–9.

10. Huang C, Wang Y, Li X, Ren L, Zhao J, Hu Y, et al. Clinical features of patients infected with 2019 novel coronavirus in Wuhan, China. Lancet. 2020 Feb 15;395(10223):497–506.

11. Yang X, Yu Y, Xu J, Shu H, Xia J, Liu H, et al. Clinical course and outcomes of critically ill patients with SARS-CoV-2 pneumonia in Wuhan, China: a single-centered, retrospective, observational study. The Lancet Respiratory Medicine. 2020 May 1;8(5):475–81.

12. Bi Q, Wu Y, Mei S, Ye C, Zou X, Zhang Z, et al. Epidemiology and transmission of COVID-19 in 391 cases and 1286 of their close contacts in Shenzhen, China: a retrospective cohort study. The Lancet Infectious Diseases. 2020 Aug;20(8):911–9.
